# The impact of COVID-19 on non-communicable disease patients in sub-Saharan African countries: systematic review

**DOI:** 10.1101/2023.10.12.23296938

**Authors:** Muluken Basa, Jan De Vries, David McDonagh, Catherine Comiskey

**Affiliations:** School of Nursing and Midwifery, Trinity College Dublin, The University of Dublin

**Keywords:** NCDs, non-communicable diseases, COVID-19 and Sub-Saharan Africa

## Abstract

**Background:** COVID-19 and its prevention measures have had a significant impact on patients with non-communicable diseases (NCDs) by disrupting routine healthcare service and increasing risk factors. These challenges were expected to be more severe in sub-Saharan Africa due to the lack of physical infrastructure and inadequate resources. The quantity of studies conducted was limited, and there was a lack of published systematic reviews in the specified region. This systematic review aimed to comprehensively assess the impact of COVID-19 on NCD patients in sub-Saharan Countries.

**Method:** This systematic review adheres to the Preferred Reporting Items for Systematic Reviews and Meta-Analyses (PRISMA) 2020 guidelines and is registered with PROSPERO (ID CRD42023387755). Extensive searches were conducted in MEDLINE, EMBASE, and CINAHL databases in November 2022, supplemented by a manual search of references, grey literature, and the WHO COVID-19 database. Inclusion criteria encompassed studies that reported on the impact of COVID-19 on NCD patients in sub-Saharan African countries, focusing on access to care, health outcomes, and factors related to NCDs. Critical appraisal of study quality was performed using the Joanna Briggs Institute (JBI) analytical cross-sectional studies critical appraisal tool. Data were extracted and synthesized, highlighting the main findings and relevant limitations.

**Findings:** This review included 26 primary studies with a cumulative sample size of 15,722 participants, conducted in six sub-Saharan African countries. Findings of these studies identified that the COVID-19 pandemic caused a disruption of 76% to 80% of regular NCDs patient care provision. The studies also identified a reduction in patient health-seeking behavior and reduced medication adherence (39.0%-63%), leading to poor treatment outcome (35.66%-55.8%). Furthermore, the pandemic and related lockdowns have been implicated in the increased prevalence of substance use, decreased physical exercise, and increased mental health problems.

**Conclusion:** This systematic review identified the complex challenges faced by NCD patients in sub-Saharan Africa during the COVID-19 pandemic. It also underlines the need to consider the indirect impact on vulnerable populations while developing pandemic prevention and control strategies for the future. The current NCD management strategies should prioritize the restoration of access to essential healthcare services while considering the multifaceted risks posed by decreased physical activity, poor dietary practices, and increased substance use.

The main limitation of this review was the study design and setting. All of the studies included in this review employed a cross-sectional design, which may result in a low quality of evidence. This study identified research conducted in only six countries among the 46 UN-classified sub-Saharan nations, which may impair the generalizability of the result.

## Introduction

Non-communicable diseases (NCDs), or chronic diseases, are illnesses of long duration with typically slow progression that cannot be transmitted from one person to another [1]. These are primarily diabetes, cancer, chronic respiratory diseases, and cardiovascular diseases, and they account for 71% of all fatalities worldwide and 80% of premature deaths in low- and middle-income countries [1]. Sub-Saharan Africa accounts for about 27% of all non-communicable disease-related deaths in Africa [2]. The COVID-19 pandemic has exacerbated the situation due to the direct and indirect impact on NCD patients [3,4]. Especially, the implementation of COVID-19 prevention and control measures, such as travel restrictions, self-isolation, quarantine, lockdown, and social distancing, have hampered routine follow-up care for people with NCDs [5,6]. WHO reported that nearly 80% of countries globally experienced substantial disruption of NCD services due to the COVID −19 pandemic [7].

The extent of disruption has varied across nations, depending on factors such as the country’s existing healthcare system and COVID-19 prevention measures [8]. In sub-Saharan countries, the impact of these disruptions has been particularly severe due to more fragile and less resilient health systems, inadequate resources, and preexisting structural inequality [9]. According to cross-sectional studies performed in Ethiopia, Nigeria, and Kenya, the COVID-19 pandemic caused 80%, 51.0%, and 42% of disruptions in NCD care, respectively [10–12]. Furthermore, these studies reported that population-level NCD screening programs had been discontinued in Ethiopia (73.3%) and Kenya (49.8%) [10,11].

People with NCDs were also found to be more vulnerable to the psychological effects of COVID-19 and required additional support due to social distancing and other preventative measures [13,14]. Moreover, studies conducted during the COVID-19 pandemic identified a significant increase in the prevalence of NCD risk factors such as cigarette smoking, alcohol drinking, inadequate physical excessive, and unhealthy diet/eating poorly [13,15,16]. An increased need for substance use treatment resulting from pandemic related stress, anxiety and social isolation has also been reported [13,17,18].

Considering these noted impacts, this systematic review is aimed at a comprehensive exploration of the existing literature on the impact of COVID-19 on non-communicable disease patients in sub-Saharan African countries. It is the first of its kind with a focus on these countries and differs from other reviews because it draws connections between multiple variables.

## Method

This systematic review made use of the Preferred Reporting Items for Systematic Reviews (PRISMA) guidelines [19] and was registered with the International Prospective Register of Ongoing Systematic Reviews (PROSPERO) (ID CRD42023387755) [20].

### Search method and selection method

The following databases were used: MEDLINE, EMBASE, and CINAHL. The search was conducted in November 2022 and included relevant medical terms and keywords with word variants for NCDs, COVID-19 and names of specific countries in the Sub-Saharan Region. An additional manual search was conducted focused on references and unpublished works of literature using Google Scholar, grey literature, and the WHO COVID-19 database. The complete search strategy is available in supplementary file, S2 File. Full search terms.

Search results were exported into Covidence, a web-based tool that manages the different steps of the procedure for conducting systematic reviews. Duplicates were removed, and the titles and abstracts of the remaining literature were screened. If deemed relevant, full texts were then reviewed for eligibility by two reviewers independently according to the pre-defined selection criteria. Any disagreements between reviewers were resolved through discussion.

### Inclusion and exclusion criteria

Studies were considered eligible for inclusion in the review only if they:

- Reported on the impact of COVID-19 on NCD patients in sub-Saharan African countries.
- Focused on one or more of the review objectives (access to care, health outcomes, risk and protective factors related to NCDs)
- Were primary research studies.
- Were published in English.
- Studies conducted in adult population (aged 18 years or older)

Studies were excluded from the review if they:

o Did not report on the impact of COVID-19 on NCD patients in sub-Saharan African countries (e.g. Studies about vaccine utilization)
o Were not published in English/ didn’t have translated version.
o Did not focus on one or more of the review objectives (access to care, health outcomes, other factors related to NCDs)
o Were not primary research studies (e.g., review articles, editorials)
o Studies conducted on different study population, for example children.

### Quality appraisal

The Joanna Briggs Institute (JBI) analytical cross sectional studies critical appraisal tool was used to assess the quality of the studies [21]. The aim of this evaluation was to evaluate a study’s methodology and ascertain how well it has addressed the possibility of bias in the research process. Eight questions are included in this checklist, which focus on methodological aspects of each study. Each question in the quality appraisal tool was given one mark, and the final score was added up, divided by eight, and multiplied by a hundred to determine a percentage. For example, if a study properly addressed 6 aspects on the checklist, it would get 6/8*100= 75%. Finally, studies were classified as high risk of Bias (score ≤49%), Moderate risk of Bias (50% - 69%) =), and low risk of Bias (above 70%), based on their respective quality score. See supplementary file, S2 Table. Risk of bias assessments. The investigators evaluated the quality of the papers independently and then met to discuss and agree on individual scoring differences.

### Data extraction method

The relevant data were identified based on the aim of the study, which includes study design, sample size and characteristics, study setting, data collection methods, and main findings related to access to care, mental health outcomes, and risk and protective factors related to non-communicable diseases (NCDs). Following study identification, data extraction was conducted using a standardized data extraction form prepared in Microsoft Word. Data extraction was done by two authors, and any discrepancies or uncertainties in the extracted data were resolved through discussion among the research team to ensure accuracy and consistency. Finally, the extracted data were checked for accuracy and completeness before data synthesis began.

### Data synthesis method

The extracted data was first arranged according to the study focus and main findings, and the emerging themes were coded. Then, the data was further organized by themes, with a separate section for each new theme and its subthemes (such as access to care and mental health outcomes, risk, and protective factors of NCDs, etc.). The synthesized data were analyzed and interpreted to identify any patterns or trends that emerge. This process involved comparing the findings across different studies to identify commonalities and differences. Additionally, any gaps or inconsistencies in the data were noted and explored further to ensure a comprehensive synthesis of the available evidence. In addition, this incorporates examining the relationships between variables (such as access to care and mental health outcomes or substance use and mental health). This systematic review also highlighted inconsistencies or gaps in the literature, and suggested areas for further research.

### Ethical approval

Since all the data was in the public domain, no ethical approval was needed for this review.

## Results

### Search results

A total of 3647 studies were imported to Covidence, with 779 duplicates. After the evaluation of the titles and abstracts of 3268 studies and the full-text screening of 255 studies, 26 met the inclusion criteria for a systematic review (see Fig. 1 for a PRISMA diagram). The diagram was developed according to the PRISMA 2020 statement: an updated guideline for reporting systematic reviews [22].

**Fig. 1.**
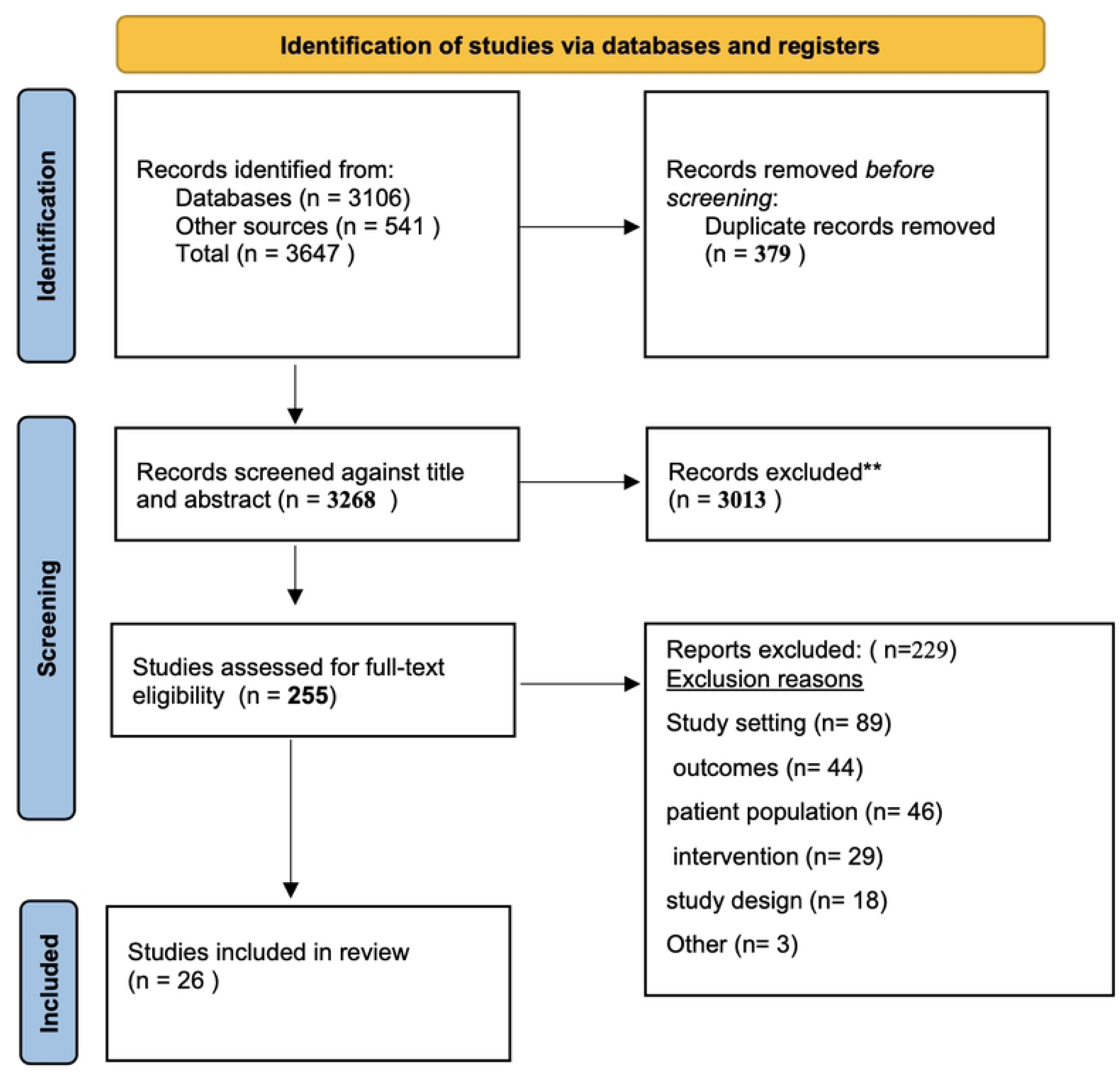
preferred reporting items for systematic reviews and meta-analyses (PRISMA) diagram.

### Characteristics of included studies

Table 1 describes characteristics of the 26 primary studies with a cumulative total of 15,722 sample size gathered from six different sub-Saharan African countries (Ethiopia, Nigeria, Rwanda, Kenya, Botswana, and Uganda). Among this, 14 studies focused on chronic disease patients, five focused on diabetes, three focused on hypertension, two focused on diseases and/ or hypertension, one focused on cancer, and one focused on cardiovascular disease. The review findings were organized into five frequently reported themes and subthemes: these are 1) disruption of follow-up care and poor health care utilization reported by 11 studies [10,11, 23,24,25,26,27,28,29,30,31]; 2) increased anxiety, stress, depression, and increased mental health problems reported by eight studies [15,32, 34,35,37,38,39,43]; 3) substance use reported by four studies [36,37,41,44]; 4) decreased physical activity and sedentary life reported by three studies [33,36,44]; and 5) increased food insecurity and poor dietary habits due to COVID-19 among NCDs patients, reported in five studies [36,40,42,43,44] Studies included in this review did not focus on the same subject matter. Some studies focused on one theme and others focus on more than two themes; for instance, (Andualem et al., 2020) examined adherence to lifestyle modifications and reported on diet, exercise, smoking, and alcohol, whereas (Abate et al., 2022) examined adherence to physical exercise recommendations and reported only on exercise compliance.

**Table 1.** Characteristics of Studies identified on the impact of covid-19 on non-communicable disease patients in sub-Saharan African countries.

| S. no. | Author, Year | Study focus | Study design/ setting | Sample size/ participants | response rate | Key findings |
| --- | --- | --- | --- | --- | --- | --- |
| 1. | (Abate et al., 2022) (33) | exercise adherence | ○ cross-sectional study<br>○ Ethiopia | ○ 576 diabetes mellitus patients | ○ 99.3% | ○ 26.4% adherence for exercise (73.6% non-adherents). |
| 2. | (Abdisa et al., 2022). (24). | medication adherence | ○ cross-sectional study<br>○ Eastern Ethiopia | ○ 402 adult hypertensive patients | • 99% | • 63% had poor antihypertensive medication adherence during pandemic with 95% CI 48.1 to 67.9. |
| 3. | (Addis et al., 2021) (34) | Psychological impact of COVID-19 | ○ cross-sectional study<br>○ Northeast Ethiopia | ○ 413 chronic disease patients | ○ 97.9% | ○ 22.8% chronic disease patients had abnormal psychological impact due to COVID-19 (95% CI: 18.6-27.1) |
| 4. | (Ajele et al., 2022b) (35) | depression and psychological well-being | ○ cross-sectional survey<br>○ Okitipupa, Nigeria | ○ 223 (age 35 to 73 years, people living with diabetes) | ○ NA | ○ psychological well-being has negative significant relationship between diabetes distress ($r = -0.42, p < .05$) and depression ($r = 0.52, p < .05$) among persons with diabetes during covid-19 pandemic. |
| 5. | (Andualem et al., 2020) (36) | Adherence to Lifestyle Modifications | ○ cross-sectional study<br>○ Northeast Ethiopia | ○ 301 hypertensive patients, | ○ 100% | ○ 23.6% adherence to lifestyle modifications.<br>○ Adherence to diet 23.6, exercise 60.1, cessation of smoking 10.1, alcohol 6.0 |
| 6. | (Awel et al., 2022) (25) | Health Seeking Behavior | ○ cross-sectional study<br>○ Southwest Ethiopia | ○ 400 chronic patients | ○ 94.8% | ○ 156 (39.0%) of them had poor health-seeking behavior. |
| 7. | (Ayalew et al., 2022) (37) | Quality of life | ○ cross-sectional study<br>○ Southern Ethiopia | ○ 633 chronic diseases | ○ 97 % | ○ significant association between different independent variables such as alcohol use in the past 3 months, common mental disorder (CMD), and QOL. |
| 8. | (Ayele et al., 2022a) (26) | Disruption of follow-up care | ○ cross-sectional survey<br>○ Northwest Ethiopia | ○ 1833 patients with common chronic disease | ○ 99% | ○ the rate of missed appointments was 12.5% (95% CI: 11.13%, 14.20%) before the pandemic, increased to 26.8% (95% CI: 24.73%, 28.82%) during the pandemic (p-value < 0.001). |
| 9. | (Ayele et al., 2022b) (27) | Treatment control | ○ cross-sectional survey design<br>○ Northwest Ethiopia | ○ 836 diabetic and/or hypertensive patients | ○ 99% | ○ Poor treatment control increased significantly from 24.81% (21.95, 27.92) prior to the COVID-19 pandemic to 35.66% (32.26, 39.20), and 34.18% (31.02, 37.49) in the second, and fourth months following the date of the first COVID-19 case detection in Ethiopia, respectively. |
| 10. | (Belete et al., 2022) (38) | Prevalence of depression | ○ cross-sectional study<br>○ Northwest Ethiopia | ○ 420 adult cancer patients | ○ 99.5% | ○ The prevalence of depression was 33.1% |
| 11. | (Ofori et al., 2021) (39) | Psychological impact | ○ cross-sectional design diabetes<br>○ Cape Coast, Ghana | ○ 157 diabetes mellitus patients aged 20 years and above. | ○ 87% | ○ About 33.8% of the study population expressed a sense of worry towards the pandemic, high level of COVID-19 specific worry, and moderate fear of isolation.<br>○ age, employment status, and the presence of other chronic diseases were significantly associated with pandemic related stress. |
| 12. | (Fentaw et al., 2022) (28) | Blood pressure control status | ○ cross-sectional study<br>○ Northeast Ethiopia | ○ 380 patients with hypertension<br>○ adults aged 18 or more | ○ 94% | ○ The prevalence of uncontrolled blood measures in patients with hypertension with a 95% CI was 55.8%(50.7%, 61.0%). |
| 13. | (Gebeyehu et al., 2022) (40) | Dietary knowledge and practice | ○ cross-sectional study.<br>○ Eastern Ethiopia | ○ 253 type 2 diabetes mellitus patients.<br>○ | ○ 91% | ○ 53.8% of type 2 diabetes mellitus patients had poor dietary practice and 52.8% found to have poor dietary knowledge. |
| 14. | (Girma et al., 2021) (32) | Stress and Coping Strategies | ○ cross-sectional study<br>○ southwest Ethiopia. | ○ 613 adults with chronic disease. | ○ 96% | ○ 68.4% of study participants were moderately stressed, 13.9% were severely stressed, and 17.8% had low levels of perceived stress. |
| 15. | (Joseph et al., 2022) (12) | Cancer Treatment | ○ cross-sectional study<br>○ Nigeria. | ○ 1,072 patients with cancer | 80.7%. | 17.3% of respondents reported disruptions to cancer care, and (51.0%) reported difficulties accessing care. |
| 16. | (Kiarie et al., 2022) (23) | Disruptions to essential health services | ○ Cross-sectional<br>○ Kenya | across the country | NA | decreases in multiple indicators, including outpatient visits (28.7%; 95% CI 16.0–43.5%), cervical cancer screening (49.8%; 20.6–57.9%), hypertension cases (10.4%; 6.0–39.4%), |
| 17. | (Maphisa and Mosarwane, 2022) (41) | Alcohol use | ○ cross-sectional study<br>○ Botswana | ○ 1318 adults with a past 12 months. | ○ NA | ○ The prevalence of alcohol use was 62.3% (95%CI= 59.7–64.9) during the second ban, and 90.4% (95%CI= 88.7–91.8) after the ban. |
| 18. | (Mekonnen et al., 2022) (11) | NCDs management services | ○ cross-sectional study<br>○ Addis Ababa, Ethiopia | ○ A total of 30 health centers were included in this study. | ○ NA | ○ The majority, 24 (80%), of the study participants perceived that the COVID-19 pandemic severely disrupted the non-communicable disease management services. |
| 19. | (Mekonnen et al., 2021) (42) | Dietary Adherence | ○ cross-sectional study<br>○ Gondar, Ethiopia | ○ 576 type 2 diabetes patients | ○ 99.3% | ○ The dietary adherence was 48.3% with [95% CI (44.1–52.4)]. |
| 20. | (Musinguzi et al., 2021) (30) | Cardiovascular Disease Prevention Program | ○ Qualitative study<br>○ Buikwe Districts in Uganda | ○ 39 community health workers (CHWs) and<br>○ 10 healthcare workers (HCW) participated in the study. | ○ NA | ○ (1) Disruption of CVD prevention services including halting screening for CVD risk factors at the community and health facility, halting sensitisation and health promotion activities at the community; (2) Reduction in patient health seeking behaviours; (3) Acute health facility staff absenteeism (4) Disruption in reporting and referral mechanisms; and (5) Disruption in supply chain. |
| 21. | (Nshimiyiryo et al., 2021) (29) | Accessing healthcare | ○ cross-sectional study,<br>○ Rwanda | ○ 220 chronic care patients | ○ 48.1% | ○ 44% reported at least one barrier to accessing healthcare.<br>○ Barriers included lack of access to emergency care (n = 50; 22.7%), lack of access to medication (n = 44; 20.0%) and skipping clinical appointments (n = 37; 16.8%). |
| 22. | (Shimels et al., 2021) (31) | Poor medication adherence | ○ cross-sectional design<br>○ Ethiopia. | ○ 409 adult with T2DM and hypertension | ○ 96.91 % | ○ poor medication adherence was 72%.<br>○ 57% reported COVID-19 pandemic has posed negative impacts on either of their follow-up visits, availability of medications, or affordability of prices. |
| 23. | (Umar et al., 2022) (10) | Cancer care | ○ cross-sectional survey<br>○ Kenya | 284 adult people with cancer in | 90 .44% | 42% of participants reported experiencing delays in their cancer care journey. |
| 24. | (Umutoniwa se et al., 2022) (43) | Food insecurity and level of depression | ○ cross-sectional study.<br>○ Rwanda. | ○ 220 non-communicable diseases, | ○ 100% | ○ 19.1% reported a reduction in daily number of meals in and 24.6% had depression. |
| 25. | (Yasin et al., 2022) (15) | Anxiety and knowledge toward its preventive measures | ○ cross-sectional study<br>○ Southeast Ethiopia. | ○ 633 adults with chronic medical illness | ○ 100% | ○ prevalence of anxiety among chronic patients in this study was 6.3% (95% confidence interval: 4.6%–8.5%) |
| 26. | (Azzouzi et al., 2022) (44) | Healthy Lifestyle Behaviors | ○ cross-sectional study<br>○ 65 countries. | ○ 3550 NCDs patients<br>○ | ○ 100% | ○ the pandemic had a significant impact on physical and mental health, changes in body weight, and tobacco consumption among people with NCDs. |

According to the JBI quality assessment score, 21 studies were found to be low risk of bias and the remaining five studies scored moderate risk of bias. Regarding response rate, 18 studies have of more than 90%, six studies did not report, and two studies have more than 80% of response rate.

### Main findings

#### Disruption of follow-up care and poor health care utilization

Disruption of follow-up care and poor healthcare utilization in sub-Saharan Africa were the most frequently reported challenges reported by ten studies included in this systematic review [10,11, 23,24,25,26,27,28,29,30,31]. Some of the reasons for this disruption include fear of contracting COVID-19, transportation challenges, and limited access to healthcare facilities due to lockdowns and restrictions. Additionally, the lack of telemedicine infrastructure and inadequate resources for virtual consultations further exacerbated the problem in the region.

The magnitude of disruption varied across the region; cross-sectional studies conducted in Nigeria [12], Kenya [23], and Ethiopia [11] reported 17.3%, 42%, and 80% of NCD care disruptions, respectively. There was also a statistically significant association between the emergence of COVID-19 and a decline in outpatient volume at NCD management services (83.3%), the closure of population-level NCD screening programs (73.3%), and the closure of disease-specific NCD clinics (76.7%) [33]. In Ethiopia, the rate of missed appointments increased from 12.5% prior to the pandemic to 26.8% during the pandemic (p < 0.001) [31].

In addition, the COVID-19 pandemic contributed to poor medication adherence among diabetic and hypertensive patients in Ethiopia, with prevalence rates of 72% and 63%, respectively [31,18]. The pandemic also exacerbated the prevalence of inadequate treatment control for NCDs in sub-Saharan Africa. In Ethiopia, the number of people with inadequate treatment control went from 24.81% (21.95% - 27.92%) before the COVID-19 pandemic to 36.69% (33.40% - 40.12% just three months after the first COVID-19 case was found [35]. A similar study in Ethiopia found that the prevalence of inadequate treatment control increased from 26.2% (22.5, 30.2) before the pandemic to 35.3% (31.6, 39.3) during the pandemic [24].

#### Impact of the COVID-19 Pandemic on Physical Activity

Importantly, four of the studies included in this systematic review reported that patients with noncommunicable diseases experienced a significant decrease in physical activity during the COVID-19 pandemic, with some variance between them [33,36,44]. As one of the four risk factors for NCDs, the decline in physical activity levels during the pandemic could potentially increase the risk of worsening NCDs. Two cross-sectional studies conducted among patients with hypertension and diabetes revealed a decrease of 30%-73.6% in the execution of a recommended exercise regimen during the pandemic [33,36]. Furthermore, this study also showed that more than 75% of participants did not adhere to recommended lifestyle changes during the pandemic [33]. Additionally, a study by Ayele et al. (2022a) demonstrated that participants with a sedentary lifestyle were significantly more likely to miss a medical appointment than their counterparts (AIRR = 1.36, 95% CI: 1.12; 1.71).

#### The impact of COVID-19 on dietary habits among NCDs

Five studies reported that the pandemic had a significant negative impact on food insecurity and dietary habits among noncommunicable diseases patients [36,40,42,43,44]. Two cross-sectional studies conducted among patients with diabetes, revealed that more than half of participants had poor dietary practices [40,42]. An analogous study conducted among adults with hypertension reported a higher proportion (74%) of nonadherence to dietary recommendation [36]. Moreover, Azzouzi (2022), reported that 52.8% of participants had insufficient dietary knowledge. In addition, a cross-sectional study conducted in three rural districts of Rwanda demonstrated that the number of daily meals consumed in households decreased by 19.1% during lockdown [43].

Several factors were identified as being associated with adherence to dietary recommendations. These included higher educational level (college and above) [AOR = 3.64, 95% CI [1.59– 8.34]], being a government employee [AOR = 2.38, 95% CI (1.13–4.99)], living in an urban area [AOR = 1.30, 95% CI (1.09–2.42)], having a smaller family size (family size of less than five) [AOR = 1.27, 95% CI (1.08–1.97)], having a good income level [AOR = 2.26, 95% CI (1.67–4.54)] [36,42].

#### Increased stress, anxiety, depression and other mental health problems

Another important impact of the COVID-19 pandemic was identified in seven studies that reported a negative impact of pandemic on the mental health and psychological well-being of noncommunicable disease patients [15,32,34,35,37,38,39,43]. Increased stress and anxiety can be a result of fear of contracting the virus, disruptions in healthcare services, social isolation, and economic hardships. The social and economic consequences of the pandemic were identified as major determinants of mental health [19, 36] with the prevalence of COVID-19-related stress and psychological effects ranging from 14.7% to 33.8% [8, 19, 26].

Risk factors for these effects include older age, unemployment, the presence of comorbid diseases, the duration of chronic illness, respiratory symptoms, and a lack of social support [19, 36]. Depression and anxiety were also shown to be relatively common among NCD patients. The prevalence of depression, ranged from 32.0% to 33.1% [8, 10, 26, 11], while the outcomes for anxiety demonstrated a huge variation across the studies with a prevalence range between 6.3% and 68.4% [9, 11, 26].

#### The impact of COVID-19 on substance use among NCD patients

Four studies included in this review showed that COVID-19 had a significant impact on substance use among NCD patients [36,37,41,44]. According to these studies, there has been a variable effect on alcohol use during the COVID-19 lockdown. While some studies have reported increased alcohol use and non-adherence to recommendations for alcohol consumption, others have noted an increase in hazardous drinking [36,37,41,44]. Sociodemographic factors, such as age and gender, political and economic factors, and the ban on alcohol sales in some countries, also played a big role in the prevalence of the problem. For example, a cross-sectional study on changes in alcohol use before, during, and after the alcohol sales ban during the COVID-19 pandemic in Botswana demonstrated a significant decrease in the prevalence of alcohol use among participants with a past 12-month drinking history, from 91.7% (95% CI: 90.1–93.1) before the second ban to 62.3% (95% CI: 59.7–64.9) during the second ban [41]. However, it returned to 90.4% (95% CI = 88.7–91.8), after the second ban. Similarly, hazardous drinking temporarily decreased by 30% during the second alcohol sales ban and increased to approximately 60% of its pre-ban levels after the ban [41].

In a cross-sectional study conducted in Ethiopia identified a statistically significant association between inadequate treatment control and hazardous drinking (AOR = 1.29, 95% CI: 1.02, 1.60). Furthermore, the smoking of tobacco was also shown to increase during the lockdown [36,37]. Political and economic factors, as well as a lack of knowledge about the risks of non-communicable diseases (NCDs) associated with smoking, played a significant role [44].

The results of this review revealed the presence of a clear relationship between the COVID-19 pandemic, mental health, and risk and protective factors for NCDs. Several studies identified a significant association between increased substance use and increased levels of anxiety and depression [36,37,41,44]. Similar associations were found between missed medical appointment and sedentary lifestyle and, poor treatment control and sedentary lifestyles, physical inactivity and substance use, and poor health-seeking behavior and substance use [25,26,27].

## Discussion

This systematic review synthesized evidence from 26 studies on the impact of COVID-19 on NCD patients in sub-Saharan African countries. This review identified that the COVID-19 pandemic impacted the delivery of a broad range of vital health services to patients with non-communicable diseases, which increased mental health problems, and risk factors for NCDs, including increased substance use, decreased physical activity, poor dietary practices, and household food insecurity in these countries.

The findings indicated a decrease in healthcare utilization, health-seeking behavior, and preventative healthcare services utilization, which could potentially lead to increased morbidity and mortality associated with NCDs [10, 11, 23–30,31]. The findings of this review were consistent with other international reviews conducted on populations in different parts of the world, including Latin American [46], Asia [47], Europe and the USA [48]. These similarities demonstrate that there was a clear pattern of disruption in access to NCD care across the world due to the pandemic. However, as our findings suggest, the problem may have been more severe in sub-Saharan countries, where the existing health system is more fragile and less resilient due to limited resources and preexisting structural inequality [9]. Factors such as socioeconomic and demographic factors, self-funding for medication costs, disruption of transportation, increased medication costs due to disruption, and poverty have been identified as contributing to the negative impact of the pandemic on NCD prevention and management in sub-Saharan countries [31]. The COVID-19 pandemic has also disrupted referral and supply chain systems, leading to limited availability and affordability of medication in Ethiopia [37,18] and Uganda [22]. Acute health facility staff absenteeism has also been observed in Uganda [35]. This could lead to a further increase in NCD-related morbidity and mortality in sub-Saharan countries, exacerbating the already existing burden of these diseases.

In addition, COVID-19 pandemic has had a significant impact on physical exercise among individuals with non-communicable diseases in sub-Saharan countries. This impact was variable across studies, with some studies finding significant decreases in adherence, and others finding small changes [33,36]. In a study by Abate et al. (2022), only 26.4% of type 2 diabetes patients were found to be adhering to exercise recommendations during the COVID-19 pandemic [33]. In contrast, Andualem (2020) found that 60.1% of hypertensive patients were adhering to exercise recommendations, although overall adherence to lifestyle modifications was low at 23.6% [36]. Similarly, studies from Canada [49] and Australia [50] reported that the pandemic was associated with a decrease in physical activity among individuals with NCDs. On the other hand, a cross-sectional study from Brazil and a systematic review of 15 studies revealed that the pandemic was not significantly associated with changes in physical activity among individuals with NCDs [6,51]. This variation is maybe due to the difference in the level and type of COVID-19 prevention measure the countries applied and the study time and setting [52]. Therefore, these diverging results suggest that the effect of the COVID-19 pandemic on physical activity among people with NCDs may depend on a number of factors, such as access to resources and support for physical activity, the specific context in which the pandemic is happening, but also individual characteristics [33,36]

Our systematic review identified that the COVID-19 pandemic had a negative impact on dietary habits among individuals with NCDs in sub-Saharan countries, with a higher prevalence of poor dietary practices and food insecurity being reported [36,40,42] While poor dietary practices are commonly reported problem in several countries, developing countries, including sub-Saharan countries, face additional challenges, such as a lack of food in households resulting from the loss of income and transportation due to the pandemic [36,40,42]. A study by Goudarzi et al. (2021) revealed that 37.4% of individuals with diabetes reported an increase in unhealthy food intake and a decrease in the intake of fruits and vegetables in Iran. In addition, this study identified that lower education levels and lower income show a significant association with the problem. Furthermore, a study conducted in Brazil demonstrated that 65.1% of individuals reported an increase in unhealthy food intake and a decrease in the intake of fruits and vegetables among hypertensive patients [53]

Moreover, the pandemic is also resulting in an increased prevalence of stress, anxiety, depression, and other mental health issues [13,15,16,38]. These findings are consistent with other studies conducted on the topic in different parts of the world [6,51]. According to WHO (2020), the negative impact of the COVID-19 pandemic on the psychological well-being of individuals with chronic diseases has not been uniform across different countries and populations. Factors such as the availability and quality of mental health care infrastructure, the degree of COVID-19 transmission, and the socio-economic status of the population have been identified as potential determinants of the impact of the pandemic on the psychological well-being of individuals with chronic diseases [13,15,16,38]. Overall, these findings highlight the importance of addressing the psychological well-being of individuals with chronic diseases during the COVID-19 pandemic [38]. In addition, research has shown that some people may use drugs or alcohol to deal with the stress and negative feelings caused by the pandemic or to pass the time during lockdowns [54]. Therefore, it is important to consider the psychological well-being of people with NCDs during pandemics, as they may be at a higher risk of experiencing anxiety and depression. Moreover, healthcare providers should be aware of the potential increase in substance use among people dealing with stress and negative emotions during this time.

‘In general, this systematic review identified the challenges faced by NCD patients in sub-Saharan Africa during the COVID-19 pandemic is very complex. It is vital health policymakers and decision-making bodies to seriously consider the indirect impact on vulnerable populations while developing pandemic prevention and control strategies for the future. The current NCD management strategies should prioritize the restoration of access to essential healthcare services while considering the multifaceted risks posed by decreased physical activity, poor dietary practices, and increased substance use.

## Conclusion

In general, our systematic review revealed that the COVID-19 pandemic impacted Sub-Saharan countries similarly to elsewhere in the world, but perhaps more so as it has exacerbated existing health inequities, especially for vulnerable populations. This highlights the need for urgent interventions to mitigate the impact of the pandemic on NCD patients in sub-Saharan African countries. Such interventions should prioritize maintaining access to essential health services and addressing social determinants of health. In addition, the findings from this review provide evidence of the relationship between COVID-19 and multiple risk factors for NCD patents, therefore, it is important for policymakers and healthcare providers to consider these factors when addressing the impact of the pandemic on NCD prevention and management. More research is needed to fully understand the long-term effects of the COVID-19 pandemic on NCDs; however, it is evident that now is the time to develop strategies to lessen these potential negative effects and to prepare for future pandemics like this one. To avoid devastating consequences for future pandemics like COVID-19, it is crucial to establish comprehensive and resilient healthcare systems that can effectively address the unique challenges posed by such outbreaks. This includes strengthening primary healthcare services, ensuring equitable access to healthcare resources, and implementing proactive measures to mitigate the negative effects on vulnerable populations. Additionally, investing in research and development of innovative technologies and interventions can further enhance our preparedness and response capabilities for future pandemics.

## Limitation of the study

The main limitation of this review was the study design and setting. All of the studies included in this review were observational in nature, which limits the ability to establish causality between NCD management strategies and their outcomes. This study included studies from six countries out of 46 UN-classified sub-Saharan nations, which may impair the generalizability of the result. Furthermore, the majority of the studies focused on urban populations, neglecting the potential differences in health behaviors and outcomes in rural areas. Therefore, caution should be exercised when applying these findings to other sub-Saharan African countries or rural communities. Future research should aim to include a more diverse range of study designs and settings to enhance the robustness and generalizability of the results.

## Contributors

Conceptualization: MB, CC, JdV, DM; Search strategy; MB, CC; Study selection: MB, CC, JdV, DM; Critical appraisal MB, CC, JdV, DM; Data Extraction; MB, CC, JdV, DM, Data analysis and interpretation: MB, CC; Drafting the manuscript: MB; Supervision; CC, JdV, DM Revision of the article: CC, JdV, DM.

## Data sharing statement

All data generated or analyzed during this study are included in this published article (and its supplementary information files).

## Data Availability

All relevant data are within the manuscript and its Supporting Information files.

## Supporting information

**S1 Fig. PRISMA diagram**

**S1 Table. Characteristics of Studies**

**S2 Table. Risk of bias assessments**

**S1 File. Review protocol**

**S2 File. Full search terms for one database.**

**S3 Table PRISMA_2020_checklist**

